# Efficacy of McKenzie Manipulative Therapy on Pain, Functional Activity and Disability for Lumbar Disc Herniation

**DOI:** 10.1101/2020.07.13.20152843

**Authors:** Mohammad Anwar Hossain, Iqbal Kabir Jahid, Md. Forhad Hossain, Zakir Uddin, Md. Feroz Kabir, K M Amran Hossain, Md. Nazmul Hassan, Lori Maria Walton

## Abstract

**Introduction:** Lumbar disc herniation (LDH) is one of the common determinations for low back pain and disability. The objectives of the study were to explore the effectiveness of McKenzie exercises and Manipulative Therapy approaches for LDH.

**Methodology:** Assessor blinded RCT carried out for 36 months at CRP. 72 subjects aged 25-50 years, clinically and radiologically diagnosed with LDH were randomly recruited and 68 found eligible. The control group received stretching exercises and Maitland mobilization, and the experimental group received McKenzie therapy for 12 sessions in 4 weeks, both groups received conventional care in addition. Pain was the primary outcome and secondary outcome was participation in functional activities and disability.

**Results:** From day 1 to 4 weeks both groups had improvement in pain, fear avoidance and bothersome (p<.05). McKenzie found superior in disability (p<.001) from 4 weeks to 6 months, in pain and disability (p<.05) from day 1 to 6 months, and in fear avoidance belief total (p<.05).

**Conclusion:** The McKenzie manipulative therapy approach reported effective for pain, disability and participating in activities for single or multiple level LDH patients from day 1 to week 4 and the treatment effect extends after 6 months.

## Introduction

In developed countries, more than 80% of the population is affected by low back pain (LBP) in some time in their life.^1,2^ The international prevalence of low back pain has been reported between 49 to 80%;^3^ Thirty-one studies have reported the prevalence of back pain in India varies from 62% in the general population to 78%, with Lumbar disc herniation (LDH) is one of the prominent causes of low back pain.^4^ LDH is defined as the localized displacement or disruption of disc material beyond the margins of the intervertebral disc space, is considered to be the most common cause of lumbosacral radiculopathy.^5^ The severity of symptoms depends upon the level of disc displacement compressing posterior or postero-lateral aspect of Lumbar spinal segments. LDH causes central low back pain and/ or radiating pain over the area of the buttocks or legs served by one or more spinal nerve roots of the lumbar vertebrae or sacrum, combined with neurologic deficits or associated symptoms of nerve root compression.^6,7^ the phenomenon can also lead to motor deficits of lumbo-sacral plexus, impairments in regular functions related to activities and livelihood.^7^

LDH is one of the most common problems confronting outpatient physical therapists. It is extensively established that herniation is a multidimensional mechanical disorder that is dependent on physical factors, lifestyle and psychosocial factors.^8^ The management of LDH depends on severity of disc displacing causing spectrum of clinical presentations^9^ and conservative treatment approach is recommended for the patients without red flags. The red flag indicates extreme pain, progressive neurological deficit and/or cauda equine syndrome. Conservative care includes a variety of pharmaceutical and non-pharmaceutical treatments such as: patient education, analgesics, rest, exercise, traction, manipulation, mobilization, manipulative therapies; clinical guideline^10^ suggests prioritizing conventional therapy as the first line of management although surgical or invasive therapies can be treatment of choice.^11,12^

The McKenzie method is widely prescribed by physical therapists to treat pain and increase flexibility for the patients having a definite mechanical characteristics of LDH symptoms.^13,14^ McKenzie Mechanical diagnosis and therapy combines exercise based on directional preferences that is intended to a “reduce derangements” and typically demonstrates one direction of repeated movement which decreases or centralizes referred symptoms, abolishes midline symptoms, along with manipulative therapy approach by the clinician, and emphasizes self-directed exercises performed by patients.^15^ McKenzie approach is evident to be effective for low back pain in contrast with pain and disability in the short term and long term, and considered as cost-effective. Hence, this is a research gap on specific concentration to lumbar disc herniation to evaluate if McKenzie manipulative therapy is effective.^15-16^ Also, there are recommendations for evaluating the therapeutic approach for the low-resource countries.^16^ The study is intended to report the effectiveness of McKenzie MDT exercises and manipulative approach for LDH patients compared to Lumbo-pelvic stretching and Maitland approach regarding outcomes of (1) pain in different functional positions, (2) fear avoidance behavior, (3) Bothersome in in functional activities and (4) low back disability index.

## Methods

The study was an assessor-blinded, randomized clinical trial (RCT), and carried out for 36 months at the Centre for the Rehabilitation of the Paralysed (CRP) in Bangladesh. The study was approved by CRP ethical review board (CRP-R&E-0401-180). The study is a fundamental feasibility study of the research project titled “Manipulative therapy for Prolapsed lumbar Intervertebral disc (PLID) patients and relation with infectious diseases: A Randomized Controlled Trail” approved by Clinical trial registry India (CTRI/2020/04/024667) the primary registry authority approved by WHO trial registry.

### Patients, Sample size calculation and Randomization

From June 2017 to December 2019, 72 patients aged 25-50 years with complain of low back pain and/or radiating pain and /or neurological symptoms towards lower limb have been primarily enrolled in the study. Then they were investigated as per inclusion criteria (diagnostic criteria). Persons having MRI and previously diagnosed as Disc herniation or Lumbar disc herniation LDH or Prolapsed Lumbar intervertebral disc (PLID) were also enrolled and screened for the second time, the persons who had no MRI were advised to perform with proper justification. Samples were enrolled in the study through hospital randomization and voluntary participation. Sixty-eight (n=68) patients complied with the eligibility criteria and were assigned after voluntary written consent, Calculated according to Miot^17^ as per MCID related to Oswestry Disability Index. Subjects were randomized either into the McKenzie group or conventional physiotherapy group with computer generated, concealed allocation. The inclusion criteria were (1) patients with a single or multiple level of lumbar disc herniation evident in Magnetic Resonance Imaging MRI, (2) positive Lasègue’s sign or cross Lasègue’s sign and (3) diagnosed as derangement syndrome 1-3 in Mechanical Diagnosis and Therapy -MDT assessment by McKenzie institute. The exclusion criteria were (1) any history of surgery for LDH, (2) co-morbidity associated with endocrine disease, osteopenia, infection or carcinoma, (3) History of fracture in the spine, ribs or upper limb within last 1 year and (4) pre-existing phobia to physiotherapy or manipulative therapy. Both groups received interventions from two outpatient settings of a hospital. Interventions were given by an experienced physiotherapist ranging 2-10 years of clinical practice experience, allocated by a random process, and a subsequent in-service training by co-researchers for the specific treatment protocol. The single assessor was blinded to the assignment and performed all assessments. The data was taken before treatment and after 12 sessions (4 weeks) of treatment in the hospital setting; a follow up was taken after 6 months of discharge by phone call or a physical visit.

### Interventions

The experimental group received McKenzie manipulative therapy for the lumbar spine. The MDT exercises included repeated movements typically include flexion in lying or standing; extension in lying or standing; and lateral movements of either side gliding or rotation and manipulative approach to lumbar spine segments.^18,19^ Patients performed those movements at therapy sessions and at home.^20^ The repeated movements of McKenzie manipulative therapy has been prescribed as 10 repetitions of directed movements, 2-3 hourly in 14 hours of a day and for 4 weeks. Manipulative therapies were performed by physiotherapists for 10-15 repetitions in a single “on/off” maneuver for 5-7 minutes for 6 sessions in 2 weeks. The control group received manual passive stretching exercise for lumbo-pelvic muscles for 5-7 repetitions per muscle with 10-15 seconds hold performed twice a day for 2 weeks and graded oscillatory mobilization in Maitland concept in 5-7 minutes, 35-40 oscillation per minutes or static segmental mobilizations in Maitland concept for 35-50 second hold for 5-7 times in lumbar spine for 6 sessions in 2 weeks In addition, both groups received analgesics and hot compression in lower back for 10 minutes for 2 weeks, stabilization exercises of lumbo-pelvic segment accompanied with a booklet indicating the proper way to do different activities and lifestyles habits for 4 weeks.^21^ All of the interventions ended up after 4 weeks from the initial day of treatment.

### Outcome measurements

Pain was the primary outcome and secondary outcome was participation in functional activities and disability. Pain was measured by the Dallas pain questionnaire (DPQ) in different activities and positions. Participation in functional activities was measured by the Fear-avoidance beliefs questionnaire (FABQ) and Sciatica Bothersome Scale (SBS) and disability was assessed with the Oswestry Low Back Disability Questionnaire (ODI). All outcome measurement tools were found to have satisfactory sensitivity and reliability.^22-26^ The outcomes were measured before intervention (day 1) and after 12 sessions (4 weeks) of intervention in the rehabilitation center setting for all the variables. A follow up was measured 6 months after discharge by phone call or a physical visit through DPQ and ODI.

### Statistical analysis

Data entry and checking quality of data was examined by an independent non-associated researcher. Data was obtained in a general linear model for paired and independent t test, and Mixed ANOVA Repeated Measures in SPSS Version 20. DPQ and ODI were analyzed utilizing a paired and independent t test for time fraction analysis and Repeated Measures ANOVA for repeated measure analysis. FABQ and SBS was analyzed utilizing a paired t-test for within group measures and independent t-test compared to baseline with a 5% level of significance. The chi-square test and independent-samples t-test were used to compare and determine the similarities of clinical baseline characteristics between the groups.

## Results

### Socio-demographic data

Sixty-eight (n=68) respondents were enrolled and randomly selected to each group. Within the control group, 3 subjects dropped out and the experimental group reported 4 subjects withdrew from the study (figure 1). In baseline assessment (table 1), the control group reported a mean age, height and weight as 38.59 ± 10.891 years, 61.38± 5.205 inches and 63.97± 8.959 Kg; and experimental group reported age at 37.71± 8.803 years, 60.50 ± 5.160 inches and 64.06± 8.180 Kg respectively. As both groups had a similar number of respondents, their occupations with service holder (Control n= 7, Experimental n=8) and housewife (Control n= 7, Experimental n=9) comprising the majority of respondents. The level of the disc herniation evident from MRI readings was reported as follows: L4/5 (Control n= 9, Experimental n=8), L5/S1 (Control n= 8, Experimental n=9) and more than one level (Control n= 14, Experimental n=13). There were no significant differences in baseline characteristics between groups (Table 1).

**Table 1:**
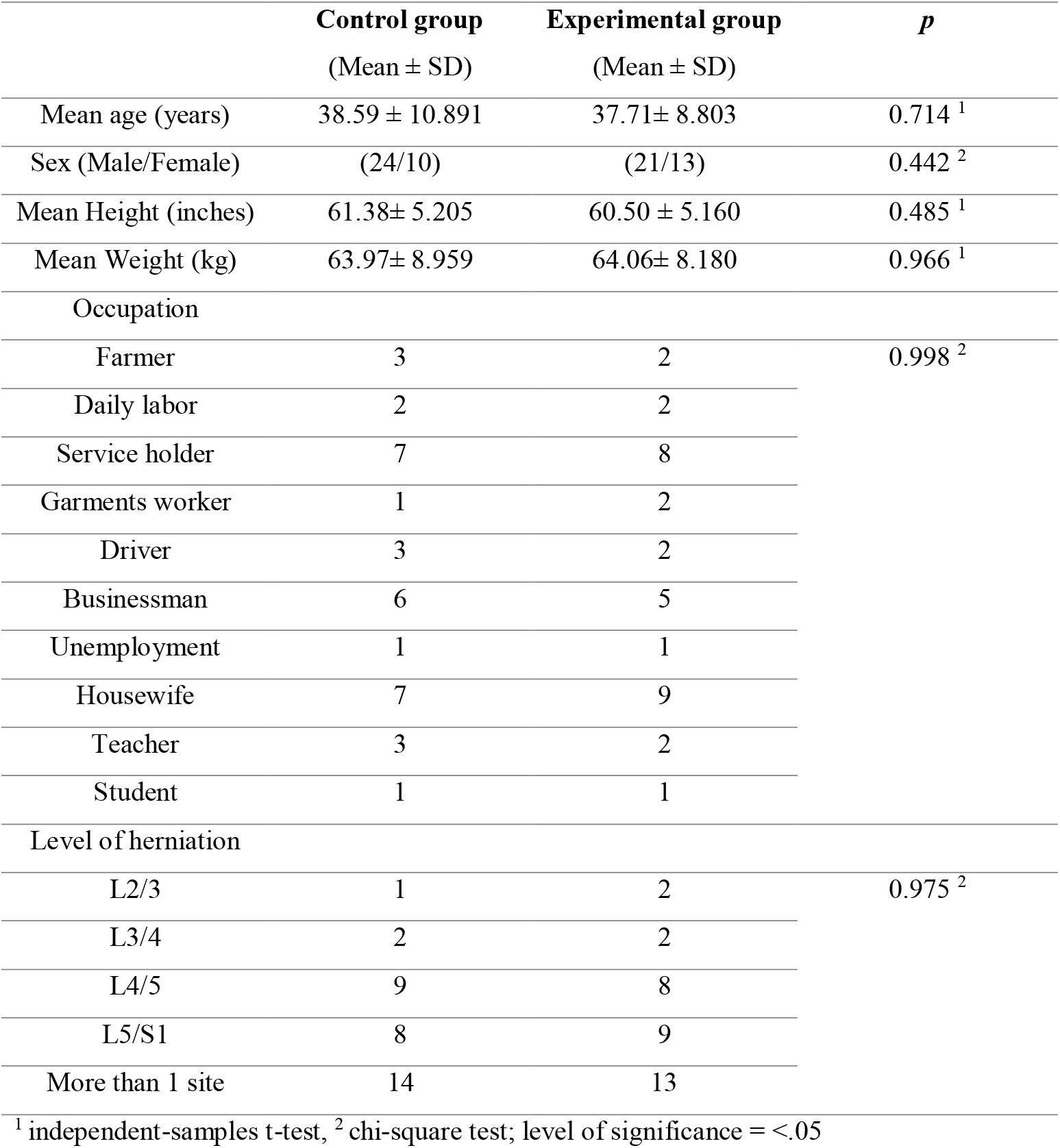
socio-demographic variables.

**Figure 1:**
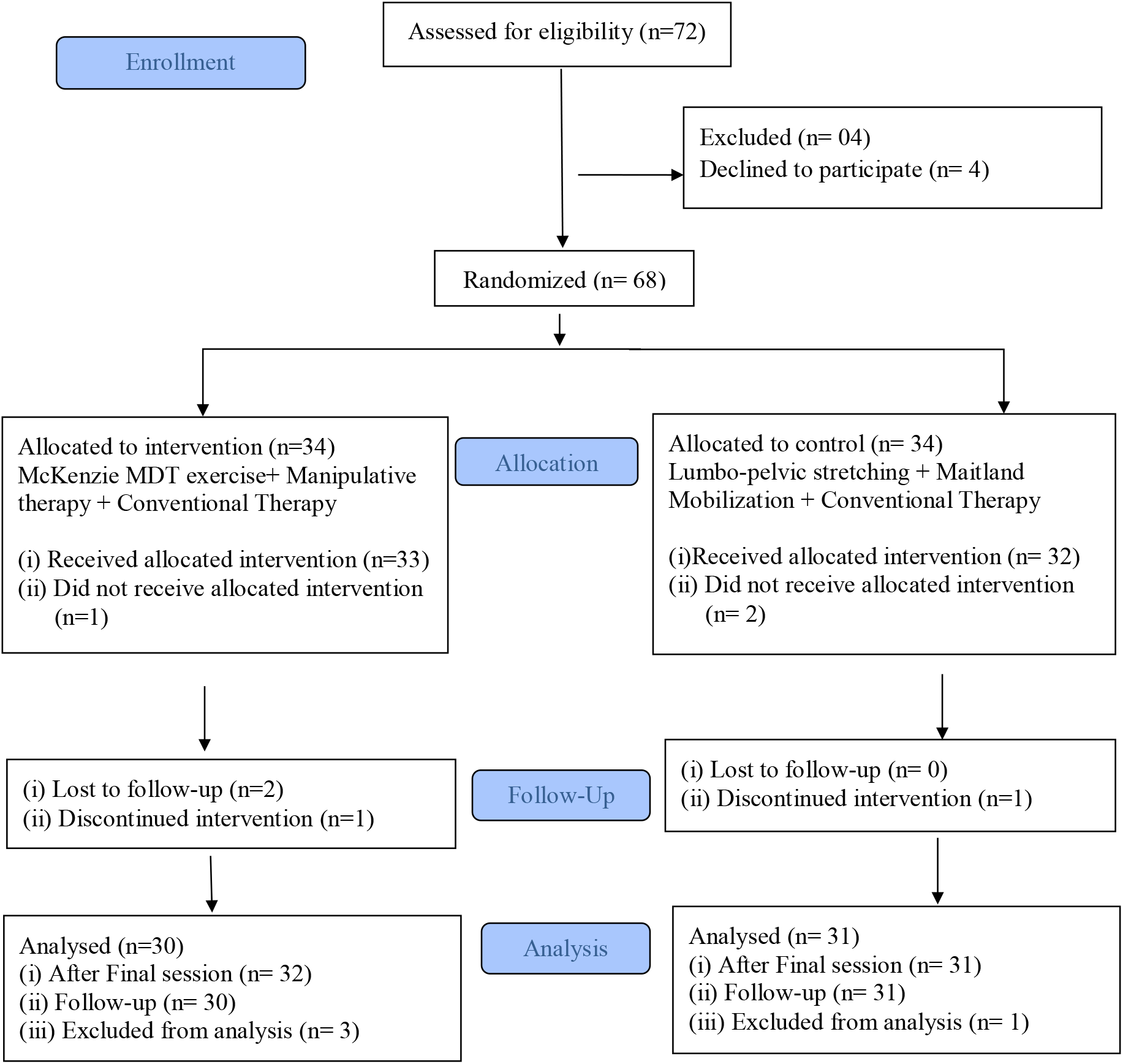
CONSORT 2010 flow diagram.

### Pain and Disability

Analysis of Dallas Pain Questionnaire (DPQ) and Oswestry Disability Index (ODI) was analyzed in three distinct statistical measures. Within group analysis of DPQ and ODI from baseline (day 1) to discharge (4 weeks) and discharge to follow up (6 months) have been conducted by paired t test (table 2) and hereby between group analysis calculated by independent t test (table 2). Changes in repeated measure from baseline (day 1) to follow up calculated with a Repeated Measures ANOVA (Table 3). Excluding the drop-out data, both control and experimental group had significant changes separately (P=<.05) in all the variables.

**Table 2:** Analysis of DPQ and ODI from baseline (day 1) to discharge (4 weeks) and discharge to follow up (6 months) separately in paired and independent t test.

| Variables | Control (day 1- 4 weeks) |  |  |  | McKenzie (day 1- 4 weeks) |  |  |  | Between group (day 1- 4 weeks) |  |  |  |
| --- | --- | --- | --- | --- | --- | --- | --- | --- | --- | --- | --- | --- |
|  | m<sup>1</sup> | 95% CI |  | p | m<sup>1</sup> | 95% CI |  | p | MD<sup>2</sup> | 95% CI |  | p |
|  |  | low | up |  |  | low | up |  |  | low | up |  |
| DPQ |  |  |  |  |  |  |  |  |  |  |  |  |
| Pain | 2.85 | 2.04 | 3.65 | .00*** | 3.77 | 2.89 | 4.65 | .00*** | -.73 | -1.5 | .13 | .09 |
| Pain at night | 2.41 | 1.48 | 3.35 | .00*** | 4.38 | 3.56 | 5.20 | .001** | -.69 | -1.5 | .13 | .09 |
| Interfere with lifestyle | 1.58 | .87 | 2.29 | .01* | 3.78 | 2.98 | 4.58 | .001** | -1.19 | -2.04 | -.33 | .001** |
| Pain severity at forward bending activity | 2.56 | 1.88 | 3.24 | .00*** | 4.38 | 3.47 | 5.28 | .00*** | -.95 | -1.88 | -.02 | .04* |
| Back Stiffness | 2.12 | 1.30 | 2.94 | .02* | 3.23 | 2.51 | 3.96 | .00*** | -1.19 | -2.07 | -.31 | .00*** |
| Interfere with Walking | 3.10 | 2.36 | 3.84 | .00*** | 4.12 | 3.29 | 4.95 | .001** | .03 | -.82 | .90 | .93 |
| Hurt when Walking | 3.16 | 2.43 | 3.88 | .01* | 2.82 | 2.09 | 3.56 | .001** | -.14 | -.98 | .70 | .74 |
| Pain keep from standing still | 2.68 | 1.96 | 3.40 | .001** | 3.76 | 2.92 | 4.60 | .01* | .172 | -.82 | 1.17 | .73 |
| Pain keep from twisting | 2.39 | 1.57 | 3.20 | .00*** | 3.09 | 2.13 | 4.04 | .00** | -.10 | -1.06 | .84 | .82 |
| Sit in upright hard chair | 2.34 | 1.58 | 3.10 | .001** | 3.00 | 2.00 | 3.99 | .001** | -.55 | -1.57 | .47 | .28 |
| Sit in soft arm chair | 2.43 | 1.68 | 3.18 | .03* | 3.27 | 2.47 | 4.06 | .02* | -1.00 | -1.99 | -.012 | .04* |
| Pain in lying | 2.19 | 1.57 | 2.81 | .00*** | 4.23 | 3.36 | 5.11 | .001** | -.80 | -1.82 | .22 | .12 |
| Pain limit normal lifestyle | 2.25 | 1.54 | 2.97 | .001** | 4.10 | 3.39 | 4.81 | .00*** | -1.58 | -2.53 | -.63 | .001** |
| Interfere with work | 2.32 | 1.56 | 3.08 | .03* | 3.58 | 2.80 | 4.36 | .00*** | -.61 | -1.57 | .34 | .20 |
| Change of workplace | 1.65 | .77 | 2.53 | .001** | 4.08 | 3.29 | 4.87 | .001** | -.65 | -1.60 | .29 | .17 |

| ODI | 12.12 | 7.48 | 16.75 | .001** | 32.77 | 25.13 | 40.41 | .00*** | -6.79 | -11.9 | -1.67 | .10 |
| --- | --- | --- | --- | --- | --- | --- | --- | --- | --- | --- | --- | --- |
| Variables | Control (4 weeks -6 months) |  |  |  | McKenzie (4 weeks -6 months) |  |  |  | Between group (4 weeks -6 months) |  |  |  |
|  | m<sup>1</sup> | 95% CI |  | p | m<sup>1</sup> | 95% CI |  | p | MD<sup>2</sup> | 95% CI |  | p |
|  |  | low | up |  |  | low | up |  |  | low | up |  |
| DPQ |  |  |  |  |  |  |  |  |  |  |  |  |
| Pain | 1.95 | 1.34 | 2.55 | .001** | 1.30 | .45 | 2.14 | .00*** | -.33 | -1.18 | .52 | .44 |
| Pain at night | .83 | .31 | 1.35 | .001** | -.34 | -1.24 | .55 | .001** | .33 | -.66 | 1.34 | .50 |
| Interfere with lifestyle | 2.41 | 1.77 | 3.05 | .02* | 1.27 | .47 | 2.06 | .001** | -.17 | -1.02 | .68 | .69 |
| Pain severity at forward bending activity | 1.44 | .80 | 2.08 | .00*** | 1.23 | .74 | 1.71 | .00*** | -.81 | -1.55 | -.06 | .03* |
| Back Stiffness | 1.52 | .72 | 2.32 | .02* | .95 | .47 | 1.42 | .00*** | -.70 | -1.48 | .08 | .07 |
| Interfere with Walking | 1.16 | .70 | 1.61 | .00*** | 1.25 | .75 | 1.74 | .001** | -.13 | -.86 | .60 | .72 |
| Hurt when Walking | 1.19 | .84 | 1.53 | .01* | 1.01 | .46 | 1.56 | .001** | .10 | -.67 | .88 | .78 |
| Pain keep from standing still | .87 | .34 | 1.40 | .001** | 1.19 | .78 | 1.59 | .02* | -.06 | -.92 | .78 | .87 |
| Pain keep from twisting | 1.13 | .48 | 1.77 | .00*** | 1.13 | .78 | 1.48 | .00** | -.31 | -1.18 | .56 | .48 |
| Sit in upright hard chair | 1.41 | .80 | 2.02 | .001** | 1.45 | .96 | 1.94 | .001** | -.67 | -1.50 | .16 | .11 |
| Sit in soft arm chair | 1.34 | .82 | 1.87 | .03* | .92 | .54 | 1.29 | .04* | -.69 | -1.67 | .28 | .16 |
| Pain in lying | 1.13 | .59 | 1.66 | .00*** | 1.16 | .69 | 1.62 | .001** | -1.03 | -1.99 | -.07 | .03* |
| Pain limit normal lifestyle | 1.66 | .96 | 2.35 | .001** | 1.25 | .77 | 1.73 | .00*** | -1.22 | -2.14 | -.29 | .01* |
| Interfere with work | 1.51 | .85 | 2.16 | .01* | 1.59 | .93 | 2.24 | .001** | -.73 | -1.49 | .03 | .06 |
| Change of workplace | 1.30 | .80 | 1.79 | .001** | 1.48 | .84 | 2.12 | .001** | -.94 | -1.79 | -.09 | .03* |
| ODI | 7.00 | 4.10 | 9.89 | .001** | 8.00 | 3.42 | 12.57 | .00*** | -8.13 | -13.25 | -3.01 | .00*** |

<sup>1</sup> mean, <sup>2</sup> mean difference, \*\*\* Significant with <.001, \*\* Significant with <.005, \* significant with P= <.05

**Table 3:**
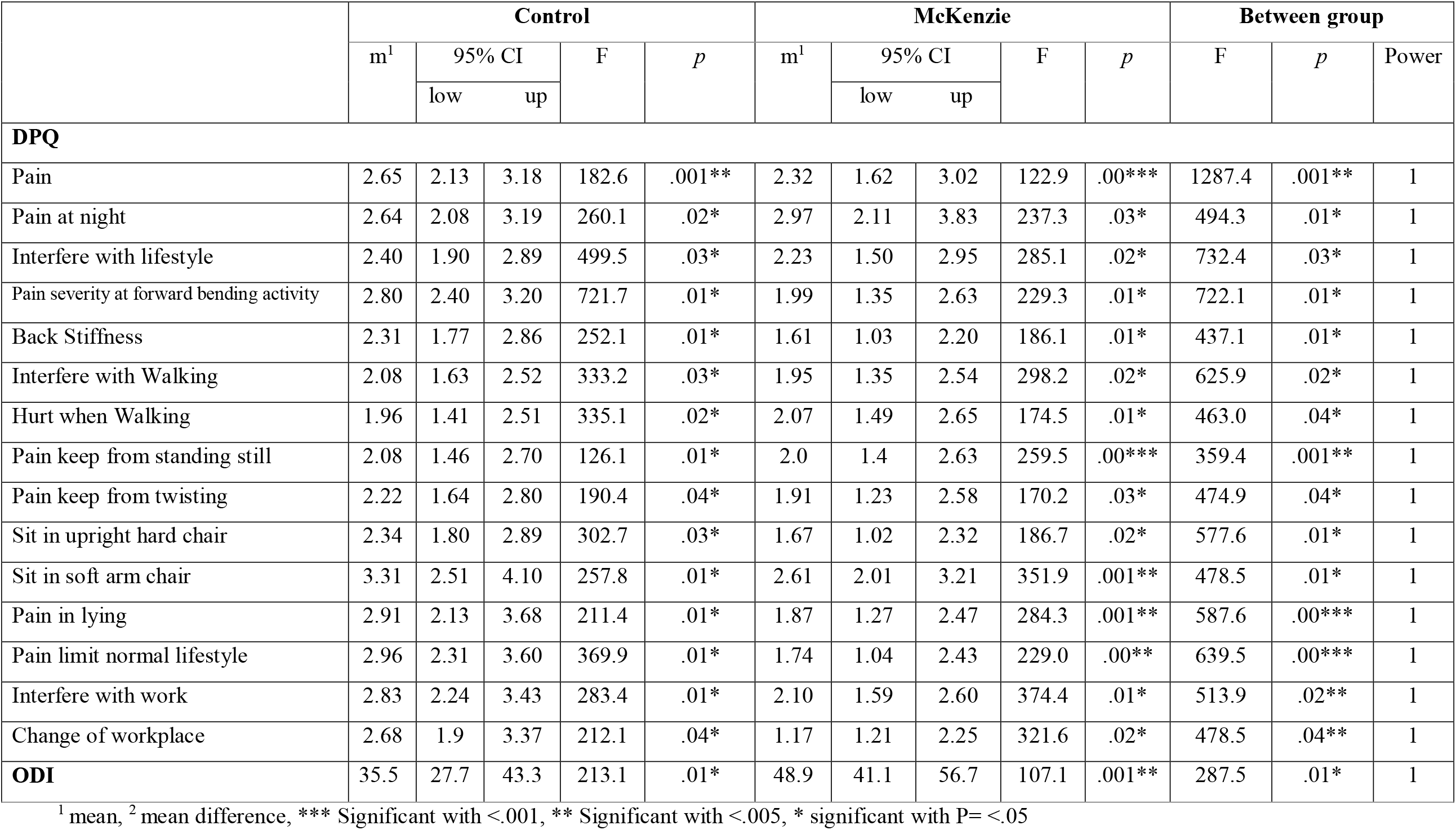
Analysis of DPQ and ODI from baseline (Day 1) to follow up (After 6 months) in Repeated measure ANOVA.

### Baseline (day 1) to discharge (4 weeks)

From baseline to discharge (Table 2) within group analysis found statistical significant changes in DPQ and ODI (p=<.05). Between group analysis found DPQ interference of lifestyle (mean difference -1.19, CI -2.4, -.33; P= <.001), Pain severity in forward bending activity (MD -.95, CI -1.88, -.02; p=<.04), Back stiffness (MD -1.19, CI -2.07, -.31; p=<.00), sit in soft arm chair (MD -1.00, CI -1.99, -.012; p=<.04) and Pain limit normal lifestyle (MD -1.58, CI -2.53, -.63; p=<.001).

### Discharge (4 weeks) to follow up (6 months)

From discharge to follow up (Table 2) experimental and control group separately found statistical significant changes in DPQ and ODI (p=<.05). Between group analysis found DPQ Pain severity in forward bending activity (MD -.81, CI -1.55, -.06; p=<.03), Pain in lying (MD -1.03, CI -1.99, -.07; <.03), Pain limit normal lifestyle (MD -1.22, CI -2.14, -.29; <.01), Change of workplace (MD -.94, CI -1.79, -.09; p=<.03) and ODI (MD -8.13, CI -13.25, - 3.01; P=<.00).

### Baseline (day 1) to Follow up (6 months)

From baseline to follow-up (Table 3) there was statistical changes in within group and between group analysis in all variables in DPQ and ODI. In control group DPQ mean varies in all the variables separately, the lowest mean was 1.96 (hurt when walking) CI (1.41,-2.51), F= 3335.1, p=<.02, highest mean was 3.31 (sit in soft arm chair), CI (2.51, 4.10), F= 257.8, P=<.01. In control group ODI represented as (mean 35.5, CI 27.7, 43.3, F= 213.1, P=<.01). In experimental group DPQ had significant changes in all variables, the lowest mean was 1.61 (back stiffness), CI 1.03, 2.20, F= 186.1, P=<.01) and highest mean was 2.97 (pain at night) CI 2.11, 3.83, F= 237.3, P=<.03. ODI in experimental group had mean 48.9, CI 41.1, 56.7. F= 107.1, P=<.001).

In between group analysis (Table 3) DPQ had significant changes (p=<.05) with F value pain 1287.4, pain at night 494.3, interfere with lifestyle 732.4, pain severity at forward bending activity 722.1, back stiffness 437.1, interfere with walking 625.9, hurt when walking 463.0, pain from standing still 359.4, pain keep from twisting 474.9, sit in upright hard chair 577.6, sit in soft arm chair 478.5, pain in lying 587.6, pain limit normal lifestyle 639.5, interfare with walking 513.9 and change of workplace 478.5 with statistical power 1. In between group analysis ODI had statistical significant change with F value 287.5 (P=<.01, power 1). The inter-quartile range (IQR) of control in the initial, discharge and follow up (Figure 2) was 34 (2.75, 20, 36.5), 23 (2, 18.0, 25) and 6.5 (13.5, 15, 20) and median found 30, 24 and 14 respectively. Also inter quartile range (IQR) of McKenzie in initial, discharge and follow up was 43 (2.75, 28, 46), 13 (1, 12, 14) and 12 (0, 7, 12) and median found 46, 14 and 6 respectively. There were also notable changes of ODI mean according to timeline in both groups and McKenzie had better remission of disability.

**Figure 2(a):**
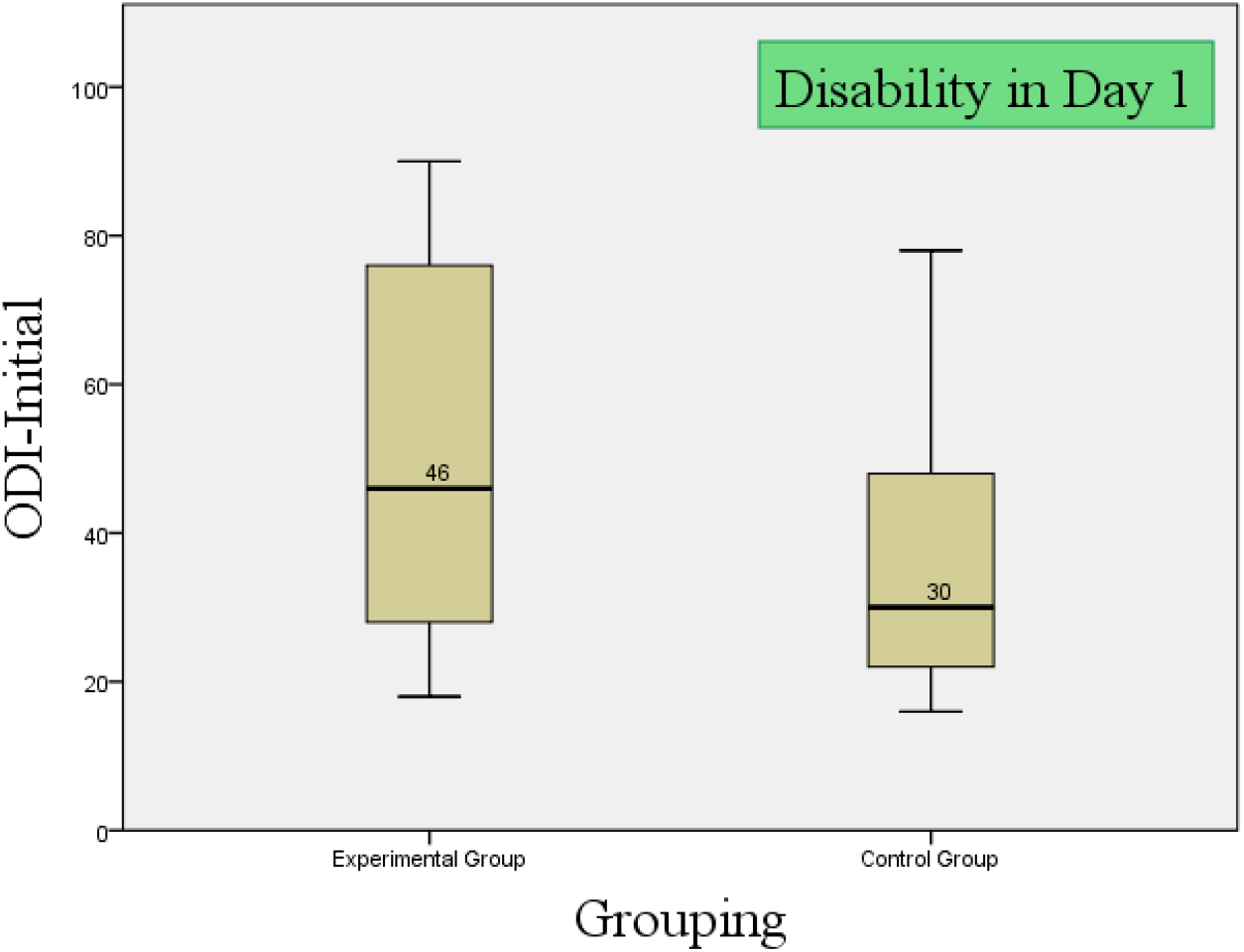
Changes of disability in ODI in day 1.

**Figure 2(b):**
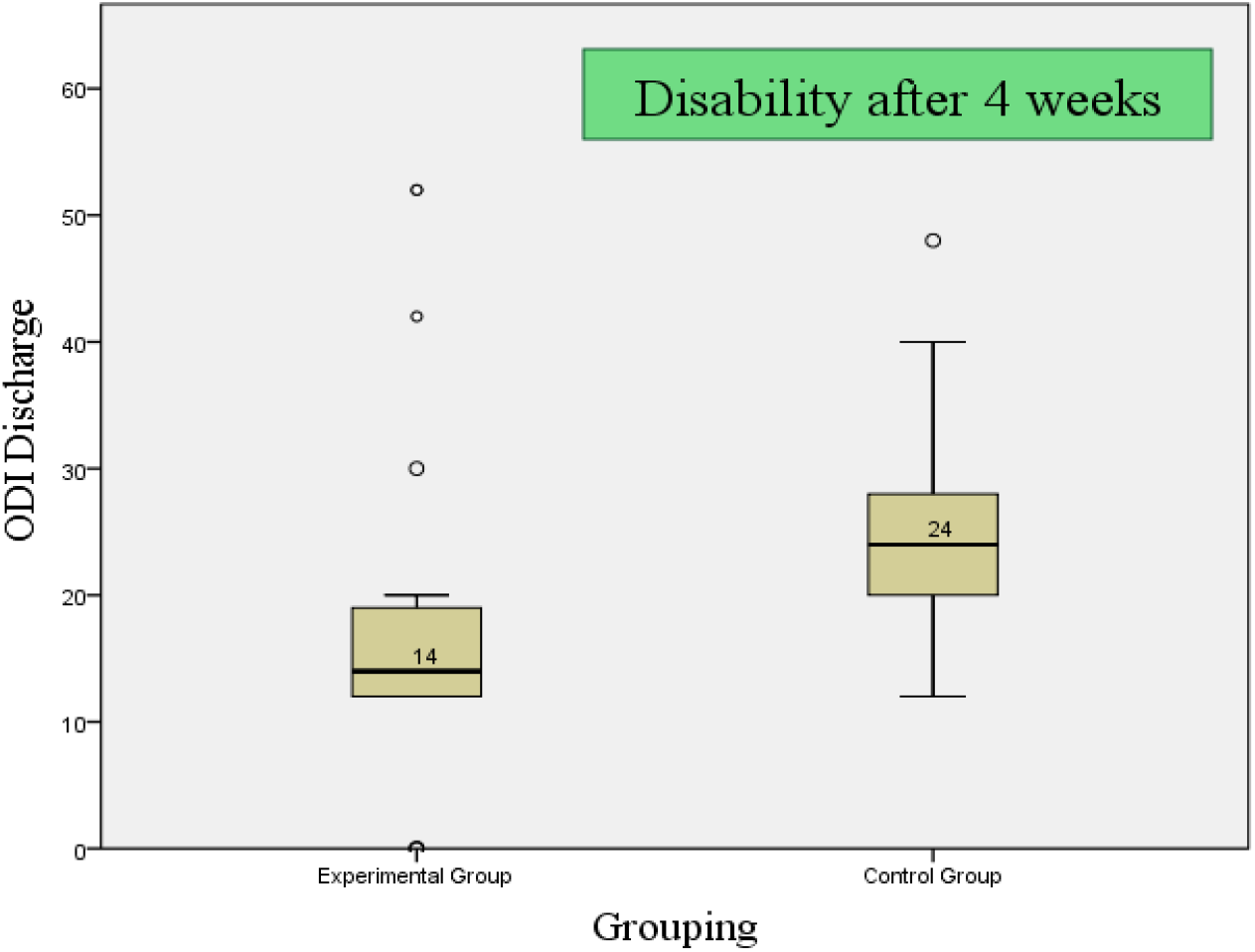
Changes of disability in ODI after week 4.

**Figure 2(C):**
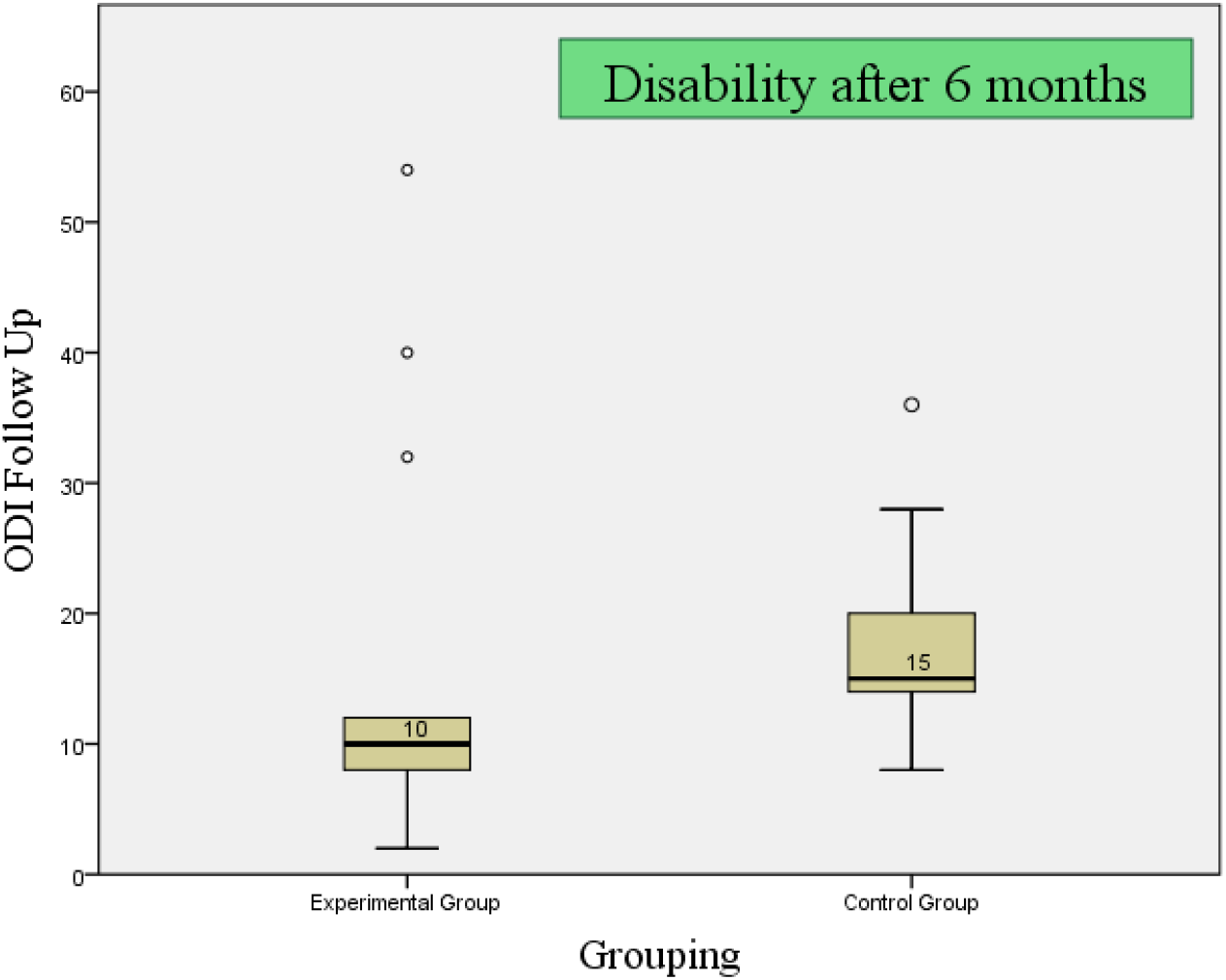
Changes of disability in ODI after 6 months.

### Fear avoidance and Bothersome in activities from baseline (day 1) to discharge (4 weeks)

From Baseline to discharge within group analysis of Fear Avoidance Belief in physical activities, work related activities reported mean differences, lower and upper value of 95% (table 4) as control 5.27, 3.99, 6.55 (p=.01), 5.78, 4.27, 7.30 (p=.01) and 16.3, 13.3, 19.2 (p=.01), and McKenzie 9.0, 7.96, 10.0 (p=.01), 16.7, 15.2, 18.1 (p=.00) and 36.0, 33.3, 38.6 (p=.00). “Bothersome due to Leg pain”, “abnormal sensation in leg”, “weakness in leg” and “leg pain in sitting” was reported with a mean difference, lower and upper value of 95% (table 3) as control 1.69, 1.31, 2.07 (p=.01), 1.63, 1.16, 2.10 (p=.000), 1.27, .690, 1.85 (p=.000) and 2.09, 1.36, 2.81 (p=.000) and McKenzie 2.16, 1.63, 2.68 (p=.01), 2.38, 1.96, 2.80 (p=.02), 2.29, 1.82, 2.75 (p=.01) and 1.38, .643, 2.13 (p=.001). The between group analysis by independent t test in FABQ reported mean difference, lower and upper value of 95% (table 3) as -1.76, -3.70, .176 (p=.074), -5.03, -7.12, -2.94 (p=.00) and -10.1, -13.8, -6.44 (p=.01), and SBI as .12, .95, .85 (p=.7), .92, 1.6, .22 (p=.02), .631, .5, .13 (p=.3) and .49, .37, 1.1 (p=.2).

**Table 4:**
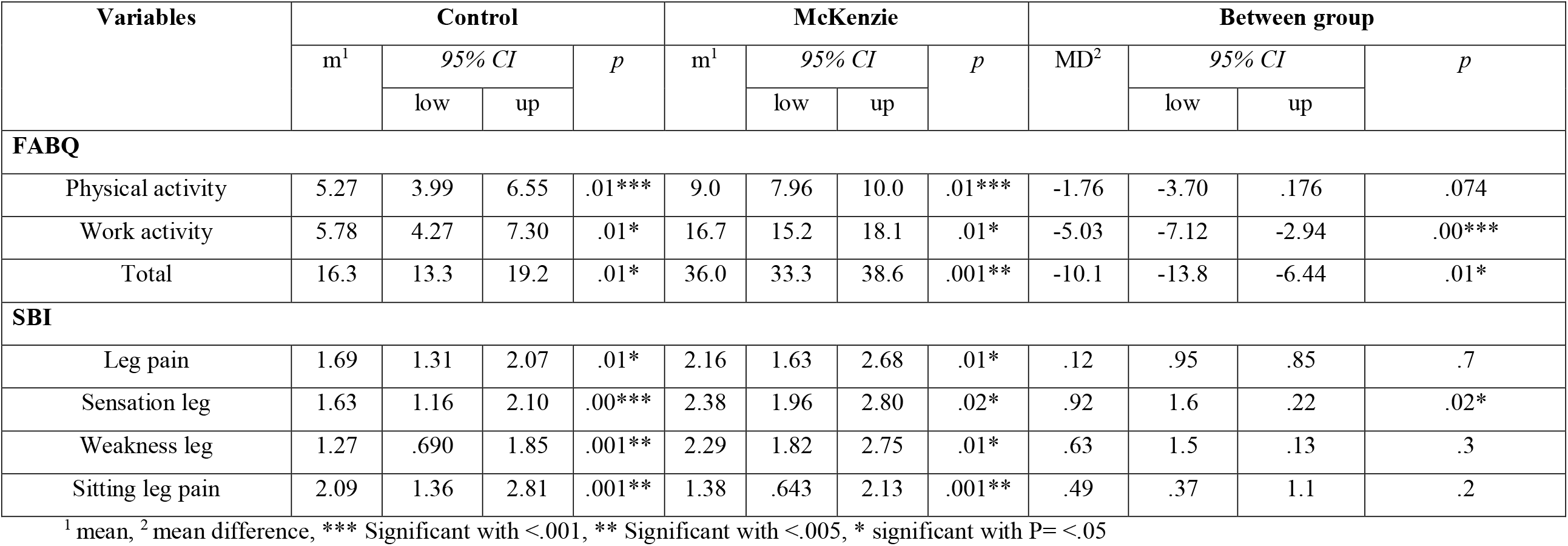
Analysis of FABQ and SBI by paired and independent t test from baseline to discharge.

| Variables | Control |  |  |  | McKenzie |  |  |  | Between group |  |  |  |
| --- | --- | --- | --- | --- | --- | --- | --- | --- | --- | --- | --- | --- |
|  | m<sup>1</sup> | 95% CI |  | p | m<sup>1</sup> | 95% CI |  | p | MD<sup>2</sup> | 95% CI |  | p |
|  |  | low | up |  |  | low | up |  |  | low | up |  |
| FABQ |  |  |  |  |  |  |  |  |  |  |  |  |
| Physical activity | 5.27 | 3.99 | 6.55 | .01*** | 9.0 | 7.96 | 10.0 | .01*** | -1.76 | -3.70 | .176 | .074 |
| Work activity | 5.78 | 4.27 | 7.30 | .01* | 16.7 | 15.2 | 18.1 | .01* | -5.03 | -7.12 | -2.94 | .00*** |
| Total | 16.3 | 13.3 | 19.2 | .01* | 36.0 | 33.3 | 38.6 | .001** | -10.1 | -13.8 | -6.44 | .01* |
| SBI |  |  |  |  |  |  |  |  |  |  |  |  |
| Leg pain | 1.69 | 1.31 | 2.07 | .01* | 2.16 | 1.63 | 2.68 | .01* | .12 | .95 | .85 | .7 |
| Sensation leg | 1.63 | 1.16 | 2.10 | .00*** | 2.38 | 1.96 | 2.80 | .02* | .92 | 1.6 | .22 | .02* |
| Weakness leg | 1.27 | .690 | 1.85 | .001** | 2.29 | 1.82 | 2.75 | .01* | .63 | 1.5 | .13 | .3 |
| Sitting leg pain | 2.09 | 1.36 | 2.81 | .001** | 1.38 | .643 | 2.13 | .001** | .49 | .37 | 1.1 | .2 |

<sup>1</sup> mean, <sup>2</sup> mean difference, \*\*\* Significant with <.001, \*\* Significant with <.005, \* significant with P= <.05

## Discussion

This research intended to explore the effectiveness of McKenzie Manipulative Therapy for LDH patients compared with a set of conventional physiotherapy treatment. The statistical analysis showed a statistically significant difference between the two groups for the ODI, with the McKenzie group having a lower score (F=107.1), which implies that the McKenzie group intervention was more effective in reducing disability than the control group (F=287.5, P=<.001) within the twelve treatment sessions, as well as follow up after six months. All the variables of Dallas pain questionnaire represented similar result. Evidence recommends^27^ using similar scales to measure disability states through physiotherapy interventions.

The control and intervention group reported similar baseline characteristics in mean age, height and weight. The occupation among groups varied, with service holder and housewife reported for the majority respondents. Two recent meta-analyses showed that subjects who were overweight or obese were at increased risk for both low back pain (LBP) and lumbar radicular pain.^23^ Abdominal obesity is defined by waist circumference and has been associated with LBP in women.^24^

As the study was conducted in the hospital setting, the priority was through the diagnosis and clinical presentations, and for concealed allocation, the groups had an insignificant similarity of baseline statistics.

Analysis of Dallas Pain Questionnaire (DPQ) and Oswestry Disability Index (ODI) has been analyzed by paired and independent t test, and repeated measure ANOVA from baseline to discharge, discharge to follow up and baseline to follow up found statistically significant difference in both group seperately. Also, between groups analysis found McKenzie concept to be superior in several parameters in several distinct timeline. From baseline to discharge McKenzie found better improvements in DPQ interference of lifestyle (mean difference - 1.19, CI -2.4, -.33; P= <.001), Pain severity in forward bending activity (MD -.95, CI -1.88, - .02; p=<.04), Back stiffness (MD -1.19, CI -2.07, -.31; p=<.00), sit in soft arm chair (MD - 1.00, CI -1.99, -.012; p=<.04) and Pain limit normal lifestyle (MD -1.58, CI -2.53, -.63; p=<.001). From discharge to follow up McKenzie group was superior in DPQ Pain severity in forward bending activity (MD -.81, CI -1.55, -.06; p=<.03), Pain in lying (MD -1.03, CI - 1.99, -.07; <.03), Pain limit normal lifestyle (MD -1.22, CI -2.14, -.29; <.01), Change of workplace (MD -.94, CI -1.79, -.09; p=<.03) and ODI (MD -8.13, CI -13.25, -3.01; P=<.00).

From baseline to follow-up McKenzie group shown better long term outcome in DPQ (P=<.05) with F value pain 1287.4, pain at night 494.3, interfere with lifestyle 732.4, pain severity at forward bending activity 722.1, back stiffness 437.1, interfere with walking 625.9, hurt when walking 463.0, pain from standing still 359.4, pain keep from twisting 474.9, sit in upright hard chair 577.6, sit in soft arm chair 478.5, pain in lying 587.6, pain limit normal lifestyle 639.5, interfere with walking 513.9 and change of workplace 478.5 with statistical power 1. In between group analysis ODI had statistical significant change with F value 287.5 (P=<.01, power 1).

However, the McKenzie group reported significantly better outcome improvement than control. The inter-quartile range (IQR) for the control was reported for the initial, discharge and follow up. Notable changes for the ODI mean was reported according to timeline in both groups, with McKenzie reporting significantly better “remission of disability” than control. Several studies suggested that McKenzie therapy was more effective than most comparative treatments at short-term follow-up in comparison with the treatments included non-steroidal anti-inflammatory drugs, educational booklet, and back massage with back care advice, strength training with therapist supervision, spinal mobilization, and general mobility exercises.^25^ Six studies were reviewed by Clare and colleagues^26^ and 1 of the 6 groups found the comparison treatment (massage/back care advice) to be more effective on both short-term and intermediate-term disability than McKenzie therapy. No other comparative treatment was more effective than McKenzie therapy at any identified point in time. Most authors focus on short-term effects of McKenzie therapy or report outcomes within 3 months of treatment but this study creates a new evidence of long term effect also. Moreover, study^27^ showing McKenzie treatment to reduce the level of disability reaching a statistical significance at 2 and 12 months follow up.

This study holds unique features that explore changes in fear avoidance beliefs in physical activities and work, and “impairments in different functional positions”. From Baseline to discharge within group analysis of Fear avoidance belief in physical activities, work related activities and total along with “Irritability due to leg pain”, abnormal sensation in leg, weakness in leg and leg pain in sitting by paired t test reported mean difference, lower and upper value of 95% found significant changes in each group separately. The between group analysis by independent t test in FABQ and SBI reported mean difference, lower and upper value of 95% found superior results in McKenzie group in FABQ activity and total, and bothersome in abnormal sensation in leg. In the study, the participants received controlled McKenzie manipulative therapy or set of conventional approach weekly three days in four weeks consecutively. Similar studies explored that^28^ six sessions over 3 weeks may bring benefits, as this study minimizes the length and proven increased frequency benefits the patient.

This study recruited 64 subjects with diagnosed LDH and allocated them, equally, in two groups of physiotherapy interventions and found significant differences in outcomes of DPQ, ODI, SBI and FABQ. One comparative randomized controlled trial reported^29^ with a 3-month follow-up period among 271 patients with chronic LBP two groups as the McKenzie therapy group (n = 134) and the other was electro physical agents group, (n = 137). In 28 sessions, significant improvement was achieved like increase in spinal motion, reduction of pain and disability within both groups but the greater improvement in the McKenzie group (p <0.05) hence, this study found improvement in pain, disability, fear avoidance and bothersome in 12 sessions. In the mentioned study, 271 samples recruited and revealed that, the McKenzie physiotherapy with different protocol like exercise or first-line care were significant, similar to this study with a minimum intervention time.

The study implied the appropriate randomization with limited resources and scarcity of samples. The assessor was blinded and the treatment provider had separate inclusion criteria and allocated to groups as per randomization process. This minimalize the potential bias and ensured masking to the patients. There was no overlap of treatment provider, hence the intervention was form of exercise which is difficult to blind to the intervention provider and patient. Patient’s participation was willing and voluntary. Because of Hospital based randomization, there was variety in demographics of the patient and in a sense despite of small sample size, the result have external validity.

The limitations of this study include smaller sample size, long duration of the study, difficulty identifying qualified subjects with specific diagnosis for inclusion factor, supported documents and eligibility criteria in timeframe of 2 years. Among the cases 5 participants (3.4%) had relapse with minimum central symptom within 6 months. Drop out analysis could improve the sample size but that was minimum in number so authors don’t considered the analysis. Calculating adverse events could improve a new dimension, the study is recommended to extend to long term prospective cohort. Future studies with multicenter, compared to surgery is recommended.

## Conclusion

The results of this study show that there is an overall statistically significant difference between the two intervention groups for the pain and disability in ODI and DPQ, but not for fear avoidance belief and bothersome in functional activities in FABQ and SBI. This is providing insight that the McKenzie method may be more effective in addition to standard physiotherapy protocol for lumbar disc herniation. However, this study was confounded by various factors, so a definitive, fully powered study is needed in the future to confirm the outcomes. This study suggests that the McKenzie Method may indeed be effective and supports the need and feasibility of a larger definitive trial in Bangladesh.

## Data Availability

accessible upon request

## Financial Support

This is a self-funded study of the authors.

## Declaration of Competing Interests

The authors declare that there are no conflicts of interest regarding the publication of this article.

### Acknowledgments

Authors acknowledges Imtiaze Ahmed and Maria Shikder for the data collection and Md. Shahoriar Ahmed and Rubayet Shafin for supporting the analysis of data.

## Data Availability

The data are available regarding this study and can be viewed upon request

## Notes

### Competing Interest Statement

The authors have declared no competing interest.

### Clinical Trial

CTRI/2020/04/024667

### Author Declarations

The study was an assessor-blinded, randomized clinical trial (RCT), and carried out for 36 months at the Centre for the Rehabilitation of the Paralysed (CRP) in Bangladesh. The study was approved by CRP ethical review board (CRP-R&E-0401-180). The study is a fundamental feasibility study of the research project titled Manipulative therapy for Prolapsed lumbar Intervertebral disc (PLID) patients and relation with infectious diseases: A Randomized Controlled Trial approved by Clinical trial registry India (CTRI/2020/04/024667) the primary registry authority approved by WHO trial registry.

## References

1. Freburger J, Holmes G, Agans R et al. The Rising Prevalence of Chronic Low Back Pain. Arch Intern Med. 2009; 169(3): 251. doi:10.1001/archinternmed.2008.543

2. An H, Thonar E, Masuda K. Biological Repair of Intervertebral Disc. Spine. 2003; 28(supplement):S86–S92. doi:10.1097/01.brs.0000076904.99434.40

3. Maniadakis N, Gray A. The economic burden of back pain in the UK. Pain. 2000; 84(1):95–103. doi:10.1016/s0304-3959(99)00187-6

4. Bindra S, Benjamin A, Sinha A. Questionnaire for low back pain in the garment industry workers. Indian J Occup Environ Med. 2013; 17(2):48. doi:10.4103/0019-5278.123162

5. Hahne A, Ford J, McMeeken J. Conservative Management of Lumbar Disc Herniation With Associated Radiculopathy. Spine. 2010; 35(11):E488–E504. doi:10.1097/brs.0b013e3181cc3f56

6. Erdogmus C, Resch K, Sabitzer R et al. Physiotherapy-Based Rehabilitation Following Disc Herniation Operation. Spine. 2007; 32(19):2041–2049. doi:10.1097/brs.0b013e318145a386

7. Luijsterburg P, Lamers L, Verhagen A et al. Cost-Effectiveness of Physical Therapy and General Practitioner Care for Sciatica. Spine. 2007; 32(18):1942–1948. doi:10.1097/brs.0b013e31813162f9

8. O’Sullivan P, Smith A, Beales D, Straker L. Association of Biopsychosocial Factors With Degree of Slump in Sitting Posture and Self-Report of Back Pain in Adolescents: A Cross-Sectional Study. Phys Ther. 2011; 91(4):470–483. doi:10.2522/ptj.20100160

9. Todd N. For debate – guidelines for the management of suspected cauda equina syndrome. Br J Neurosurg. 2010; 24(4):387–390. doi:10.3109/02688697.2010.500419

10. Traeger A, Buchbinder R, Elshaug A, Croft P, Maher C. Care for low back pain: can health systems deliver?. Bull World Health Organ. 2019; 97(6):423–433. doi:10.2471/blt.18.226050

11. Albert H, Manniche C. The Efficacy of Systematic Active Conservative Treatment for Patients With Severe Sciatica. Spine. 2012; 37(7):531–542. doi:10.1097/brs.0b013e31821ace7f.

12. Qaseem A, Wilt T, McLean R, Forciea M. Noninvasive Treatments for Acute, Subacute, and Chronic Low Back Pain: A Clinical Practice Guideline From the American College of Physicians. Ann Intern Med. 2017; 166(7):514. doi:10.7326/m16-2367

13. Kumar S, Dunsford, Clarke. Integrating evidence into practice: use of McKenzie-based treatment for mechanical low back pain. J Multidiscip Healthc. 2011:393. doi:10.2147/jmdh.s24733.

14. Lawrence D, Meeker W, Branson R et al. Chiropractic Management of Low Back Pain and Low Back-Related Leg Complaints: A Literature Synthesis. J Manipulative Physiol Ther. 2008; 31(9):659–674. doi:10.1016/j.jmpt.2008.10.007

15. Halliday M, Garcia A, Amorim A et al. Treatment Effect Sizes of Mechanical Diagnosis and Therapy for Pain and Disability in Patients With Low Back Pain: A Systematic Review. Journal of Orthopaedic & Sports Physical Therapy. 2019; 49(4):219–229. doi:10.2519/jospt.2019.8734

16. Namnaqani FI, Mashabi AS, Yaseen KM, Alshehri MA. The effectiveness of McKenzie method compared to manual therapy for treating chronic low back pain: a systematic review. J Musculoskelet Neuronal Interact. 2019; 19(4):492–499.

17. Miot HA. Sample size in clinical and experimental trials. J Vasc Bras. 2011; 10(4):275–8.

18. McKenzie R, May S. The lumbar spine: mechanical diagnosis and therapy. Orthopedic Physical Therapy; 2003 Jun 1.

19. Lam O, Strenger D, Chan-Fee M, Pham P, Preuss R, Robbins S. Effectiveness of the McKenzie Method of Mechanical Diagnosis and Therapy for Treating Low Back Pain: Literature Review With Meta-analysis. Journal of Orthopaedic & Sports Physical Therapy. 2018; 48(6):476–490. doi:10.2519/jospt.2018.7562

20. Menon A, Korner-Bitensky N, Kastner M, McKibbon K, Straus S. Strategies for rehabilitation professionals to move evidence-based knowledge into practice: A systematic review. J Rehabil Med. 2009; 41(13):1024–1032. doi:10.2340/16501977-0451

21. Turk D, Burwinkle T. Cognitive-Behavioral Perspective on Chronic Pain Patients. Crit Rev Phys Rehabil Med. 2006; 18(1):1–38. doi:10.1615/critrevphysrehabilmed.v18.i1.10

22. Marty M, Courvoisier D, Foltz V et al. How much does the Dallas Pain Questionnaire score have to improve to indicate that patients with chronic low back pain feel better or well?. European Spine Journal. 2015; 25(1):304–309. doi:10.1007/s00586-015-3957-3

23. Shiri R, Karppinen J, Leino-Arjas P, Solovieva S, Viikari-Juntura E. The Association Between Obesity and Low Back Pain: A Meta-Analysis. Am J Epidemiol. 2009; 171(2):135–154. doi:10.1093/aje/kwp356

24. Han T, Schouten J, Lean M, Seidell J. The prevalence of low back pain and associations with body fatness, fat distribution and height. Int J Obes. 1997; 21(7):600–607. doi:10.1038/sj.ijo.0800448

25. Busanich BM, Verscheure SD. Does McKenzie therapy improve outcomes for back pain?. J Athl Train. 2006; 41(1):117–119.

26. Clare H, Adams R, Maher C. A systematic review of efficacy of McKenzie therapy for spinal pain. Australian Journal of Physiotherapy. 2004; 50(4):209–216. doi:10.1016/s0004-9514(14)60110-0

27. Murtezani A, Govori V, Meka V, Ibraimi Z, Rrecaj S, Gashi S. A comparison of McKenzie therapy with electrophysical agents for the treatment of work related low back pain: A randomized controlled trial. J Back Musculoskelet Rehabil. 2015; 28(2):247–253. doi:10.3233/bmr-140511

28. Petersen T, Larsen K, Nordsteen J, Olsen S, Fournier G, Jacobsen S. The McKenzie Method Compared With Manipulation When Used Adjunctive to Information and Advice in Low Back Pain Patients Presenting With Centralization or Peripheralization. Spine. 2011; 36(24):1999–2010. doi:10.1097/brs.0b013e318201ee8e

29. Machado L, Maher C, Herbert R, Clare H, McAuley J. The effectiveness of the McKenzie method in addition to first-line care for acute low back pain: a randomized controlled trial. BMC Med. 2010; 8(1). doi:10.1186/1741-7015-8-10

